# Evaluation of the ELITe InGenius PCR assay compared with immunofluorescence for *Pneumocystis jirovecii* detection in respiratory samples

**DOI:** 10.1101/2025.11.18.25340486

**Authors:** Isabelle Dätwyler, Herbert Kuster, Michael A. Greiner, Patrick M. Meyer-Sauteur, Urs W. Karrer, Fabian Tschumi, Christoph Berger, Bhavya Chakrakodi, Philipp Bosshard, Annelies S. Zinkernagel, Roger D. Kouyos, Huldrych F. Günthard, Silvio D. Brugger

## Abstract

**Introduction:** *Pneumocystis jirovecii* (PJ) causes pneumonia primarily in immunocompromised individuals. Although direct immunofluorescence (IF) remains the diagnostic standard, PCR assays are increasingly used because of higher sensitivity and reduced observer dependency. This study evaluates the ELITe InGenius PJ PCR assay compared with IF for PJ detection in respiratory specimens.

**Material and Methods:** Respiratory samples submitted for IF-based PJ testing at the University Hospital Zurich over a 19-month period were retrospectively analyzed using the ELITe InGenius PCR assay. Diagnostic accuracy was assessed using IF as the reference method, and performance was evaluated by receiver operating characteristic (ROC) analysis.

**Results:** A total of 222 samples from 213 patients were included (70 sputum, 152 bronchoalveolar lavage). PCR and IF results were concordant in 160 (72.1%) specimens, including 18 positives and 142 negatives. Sixty-two (27.9%) samples were discrepant: six IF-positive/PCR-negative and 56 IF-negative/PCR-positive. IF reexamination was possible for IF-positive/PCR-negative and IF-negative/PCR-high-positive samples (>100,000 copies/ml or Ct <29), resulting in revised IF findings in 3/3 and 2/8 cases, respectively. Three IF-negative/PCR-positive cases were clinically confirmed as true infections. ROC analysis identified an optimal PCR threshold of 6,233 copies/ml, yielding 70.8% sensitivity and 88.3% specificity, improving to 82.6% and 89.5% after reexamination.

**Discussion:** These findings support the higher sensitivity of PCR relative to IF for direct PJ detection. Clinical confirmation of IF-negative/PCR-positive cases and known limitations of IF underscore the diagnostic value of the ELITe InGenius assay. Its performance supports its use in routine diagnostics to improve early detection and guide timely treatment.

## Introduction

*Pneumocystis jirovecii* (PJ) is an atypical, unicellular fungus with tropism for lung tissue causing pneumonia almost exclusively in immunocompromised patients^1–5^. Severe immunodeficiency remains the most essential host factor for *Pneumocystis jirovecii* pneumonia (PJP). Over the last two decades, the ratio of affected non-HIV-to HIV patients steadily increased in resource rich countries^6–9^. This shift is strongly linked to the widespread use of antiretroviral therapy (ART), which preserves cellular immunity and reduces opportunistic infections and mortality in HIV-positive patients^10–14^. Additionally, the increasing number of patients undergoing immunosuppressing or -modulating therapy for conditions such as solid organ transplantation, neoplastic, rheumatological and inflammatory diseases, has resulted in non-HIV patients representing the majority of PJP patients^15–18^.

This changing epidemiology of PJP has introduced new challenges in its identification and management^9^. PJP typically manifests as a non-specific pneumonia with symptoms including fever, dyspnea and a non-productive cough, and is associated with a high mortality rate ranging from 27 to 55%^1–3, 19, 20^. Compared to people with HIV (PWH), non-HIV patients often experience a more acute onset and critical disease course, with higher rates of respiratory failure, ICU admission, and mortality^9, 19^. In contrast, PWH tend to exhibit a more prolonged disease progression and higher fungal loads. Radiological findings, including ground-glass opacities and, in more advanced stages, a denser mosaic pattern can support the clinical suspicion of PJP^21, 22^.

Routine diagnostics do not rely on growing PJ in culture, due to its very demanding nature^2, 4, 5, 23^. Instead, microscopy or PCR analysis from respiratory specimens are used for rapid direct pathogen detection^2, 4, 23, 24^. Microscopy with conventional and immunofluorescent staining methods allows the visualization of cysts and trophozoites, with direct immunofluorescence (IF) demonstrating a more robust performance than conventional microscopy and is still considered the current diagnostic gold standard^2, 23^. In the last decades, PCR-based methods have gained importance due to their high sensitivity (97–99% in three meta-analyses), consistent performance regardless of HIV status, and reduced reliance on investigator experience^23, 25^. While it is essential to integrate pretest probability from clinical and radiological findings, particularly for the distinction between colonization and active infection, these promising PCR performance reports support a more time-efficient and sensitive PJP diagnosis.

We evaluated the ELITe InGenius PJ PCR assay as a not yet assessed platform in published studies. This study aimed to compare its performance with IF, as the reference standard, to support its implementation in routine diagnostics. It also provides a comprehensive overview to guide interpretation and establish a reliable threshold for clinical decision-making in PJP diagnosis.

## Material and methods

### Study design and population

This study includes all samples submitted for PJ testing by IF at the University Hospital Zurich (USZ) between 13.03.2023 and 22.10.2024. The patient specimens originated from multiple hospitals and laboratories across the German-speaking part of Switzerland. In this study, all samples were retrospectively analyzed using ELITe InGenius PJ PCR assay to evaluate its suitability for implementation in routine diagnostic.

### Sample processing and immunofluorescence assay

Sample preparation varied depending on the sample type following the internal protocol^26^. For bronchoalveolar lavage (BAL) samples, 0.2 to 0.5 ml were cytocentrifuged onto polytetrafluoroethylene (PTFE)-printed microscope slides using a Cytospin 4 cytocentrifuge. For viscous samples, 0.5 to 5ml of sputum, tracheal or bronchial secretions, or 1 to 5ml of mucoid BAL samples, were mixed with 6 to 7 ml of a solution containing 20 mM dithiothreitol and 20 mM ethylenediamine-tetraacetic acid (EDTA) and incubated at 37°C for 5 to 10 minutes. After dilution with phosphate-buffered saline (PBS), the samples were centrifuged, the supernatant discarded, and the sediment resuspended in a small volume of PBS (depending on the pellet size). Two to three drops of the suspension were applied to PTFE-printed slides and allowed to air dry.

Three slides were prepared from each sample for microscopic analysis^27, 28^. Two of the slides were stained for conventional microscopy with Toluidine blue and Giemsa stain, respectively. The last slide was processed by direct immunofluorescence using either MONOFLUO™ Pneumocystis jirovecii IFA (Bio-Rad) for 148 samples or Merifluor® Pneumocystis (Meridian Bioscience) for 74 samples, according to the instructions of the manufacturers. Discrepant samples with sufficient material were reexamined using Bio-Rad staining for a second confirmatory IF.

### Real-time PCR assay

Following microscopy, PCR-testing was performed on the ELITe InGenius instrument using the corresponding Pneumocystis ELITe MGB® Kit. This fully automated system extracts nucleic acids and amplifies the mitochondrial large subunit (mtLSU) rRNA gene specific to PJ. Samples were classified as negative if no DNA was detected or if the concentration was below the assay’s limit of detection (LoD), defined as 97copies/ml according to the manufacturer.

### Statistical Analysis

To evaluate the diagnostic performance of the InGenius PCR assay relative to the IF assay, receiver operating characteristic (ROC) analysis was performed. Sensitivity and specificity were calculated across a range of thresholds to identify clinically relevant cut-offs. The optimal threshold was defined as the value maximizing Youden’s index (Specificity + Sensitivity - 1). Data analysis was conducted using R (version 4.4.3) in RStudio (version 2024.12.1) including the package pROC^29, 30^. To further contextualize the findings, a sensitivity analysis was performed incorporating detailed patient histories and additional data processing to account for potentially false-positive initial IF results (see Results).

### Ethical considerations

The ethics committee of the Canton Zurich reviewed the study protocol (Kantonale Ethikkommission Zurich BASEC ID Req-2025-01064) and concluded that the study did not meet the criteria for requiring ethical approval within its remit. All research was carried out in accordance with Good Clinical Practice standards.

## Results

### Patient and sample characteristics

A total of 222 respiratory samples from 213 patients were retrospectively analyzed using quantitative real-time ELITe InGenius PJ-PCR and results compared to prior PJ detection by IF and conventional microscopy (Table 1).

**Table 1:** Patient and sample characteristics (*n*=213 patients, 222 samples)

| **Number of patients** | 213 (208 adults + 5 children) |
| --- | --- |
| **Age\*** | Median: 62 years (range: 13 days – 89 years) |
| **Sex: male/female** | 135/78 |
| **Year of sample collection: 2023/2024** | 190/32 |

*\*The age calculation refers to the year in which the sample was taken. For patients with multiple samples, age was recorded only once, as all samples were collected within the same year.*

### Primary diagnostic performance

24 (10.8%) samples tested positive by microscopy. Of these 23, 9 and 6 were positive by IF, Toluidine blue and Giemsa, respectively. In one case, only Toluidine blue staining was positive and IF negative. It was classified as microscopy-positive and included in the subsequent analysis. For clarity, results incorporating this sample are referred to as “microscopy” rather than “IF”.

Quantitative PCR detected PJ DNA in 74 (33.3%) samples with a mean concentration of 449’108 cp/ml (range: 0–31’622’777 cp/ml). The initial, unaltered results from both diagnostic methods (microscopy and PCR) were referred to as “Primary data”. Within this dataset, 18 (8.1%) samples tested positive by both PCR and microscopy, 56 (25.2%) were only PCR positive and six (2.7%) samples were only IF positive. The latter were labelled as potentially false-negative PCR using microscopy as the reference standard. In one sample from this group, a single cyst was observed during the initial IF examination but could not be confirmed by a second observer a few days later, highlighting the variability and observer-dependency of IF as a diagnostic standard.

### IF reexamination of discrepant samples

Among the six IF-positive/PCR-negative samples and 13 of the IF-negative/PCR-positive samples with high PCR positivity (defined as >100’000 copies/ml or Ct value <29), reexamination by IF was performed when sufficient material was available (Figure 1). This was possible in 3/6 IF-positive/PCR-negative and 8/13 IF-negative/PCR-positive samples. The reexamination was conducted using the Bio-Rad IF staining reagent, selected for its superior diagnostic performance in our study and reported lower specificity of Merifluor in comparison to other staining agents^31–33^.

**Figure 1:**
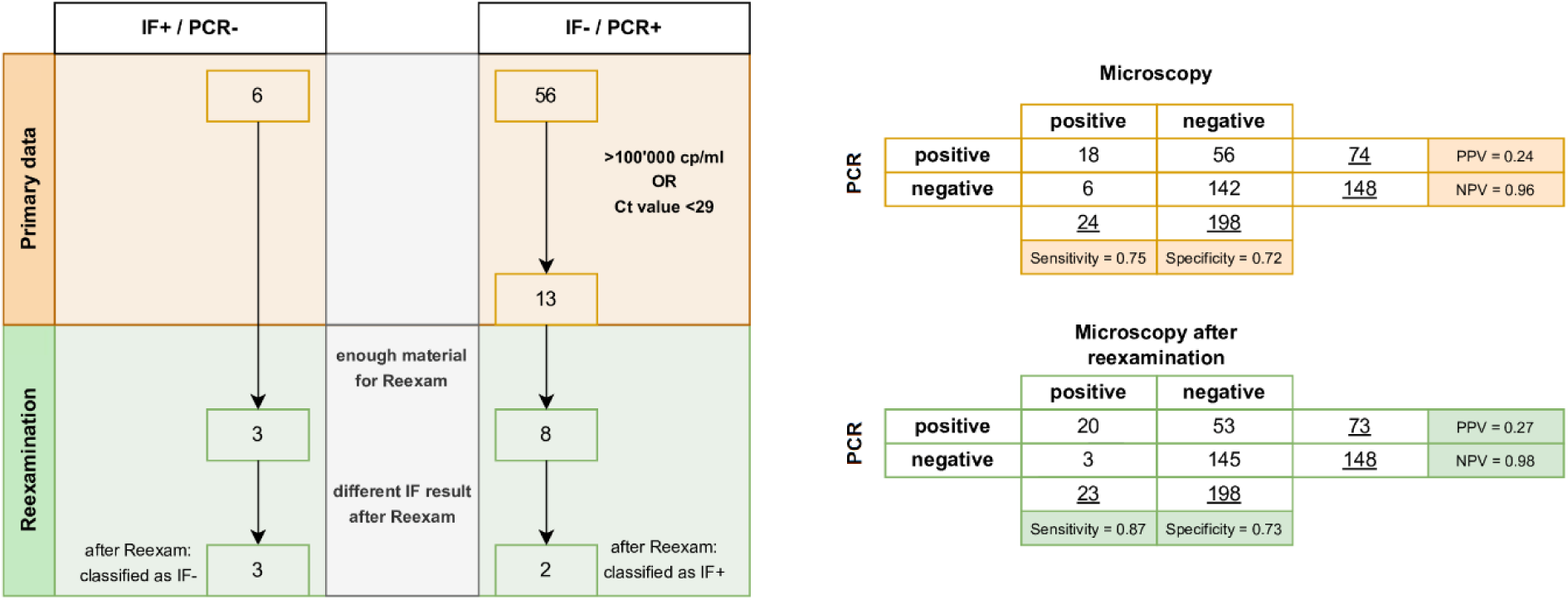
**a)** Flowchart showing data processing of discrepant results in microscopy and PCR analysis. The unadjusted primary data (orange) includes six IF+/PCR- and 56 IF-/PCR+ samples, 13 of which had high PCR positivity. IF reexamination (green) covered three false-negative and eight false-positive samples, with altered results in all three IF+/PCR- and two IF-/PCR+ cases. **b)** Contingency tables comparing PCR and microscopy results before and after reexamination, including diagnostic performance values.

In all three reexamined IF-positive/PCR-negative samples, the initial positive IF result could not be reproduced. This suggests a true-negative result further supported by a negative result in a second confirmatory PCR. In 2/8 reexamined IF-negative/PCR-positive samples, cysts were found confirming them as true positives. Based on these findings, the dataset was modified accordingly and referred to as the “Reexamination” in the following analyses.

### Diagnostic performance: IF vs. ELITe InGenius PJ PCR assay

The diagnostic performance of the ELITe InGenius PJ PCR assay was evaluated against IF as the reference standard (Figure 2). The area under the ROC curve (AUC) in the “Primary data” was 0.82, and when using the assay’s limit of detection (LoD; 97 cp/ml) as the cut-off, it demonstrated a sensitivity of 75.0% and specificity of 71.7%. Following reexamination, the AUC increases to 0.9, sensitivity to 87.0% and specificity to 73.2%.

**Figure 2:**
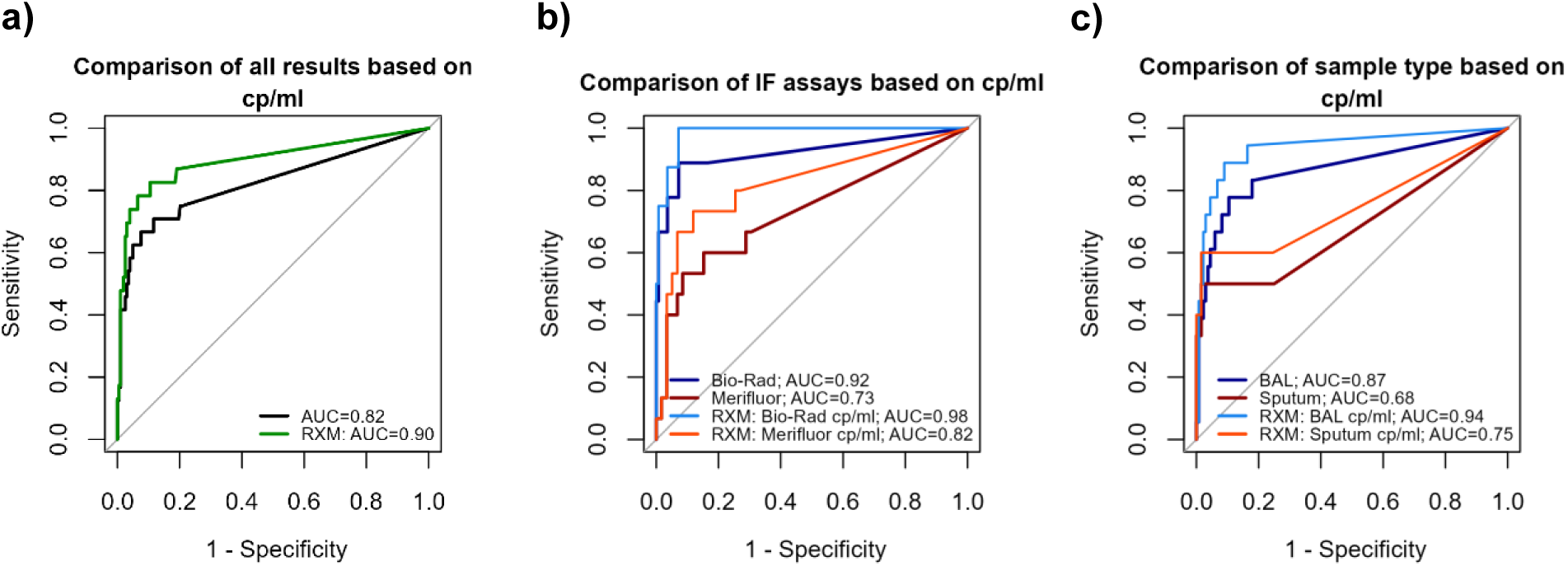
ROC curves evaluating PCR performance against microscopy as the reference standard including the “Primary data” and “Reexamination” (RXM) data sets. **a)** All samples; **b)** IF assay stratified by supplier (Bio-Rad vs. Merifluor); **c)** IF assay stratified by sample type (BAL vs. sputum)

ROC analysis identified an optimal threshold of 6233 cp/ml in both datasets, maximizing the sum of sensitivity and specificity. At this cut-off, the “Primary data” showed a sensitivity of 70.8% and specificity of 88.3%. After reexamination, these values improved to 82.6% and 89.4%, respectively. In both datasets, microscopy-positive samples predominantly exceed a copy number of 6233 cp/ml, while microscopy-negative samples more often show lower values (Figure 3).

**Figure 3:**
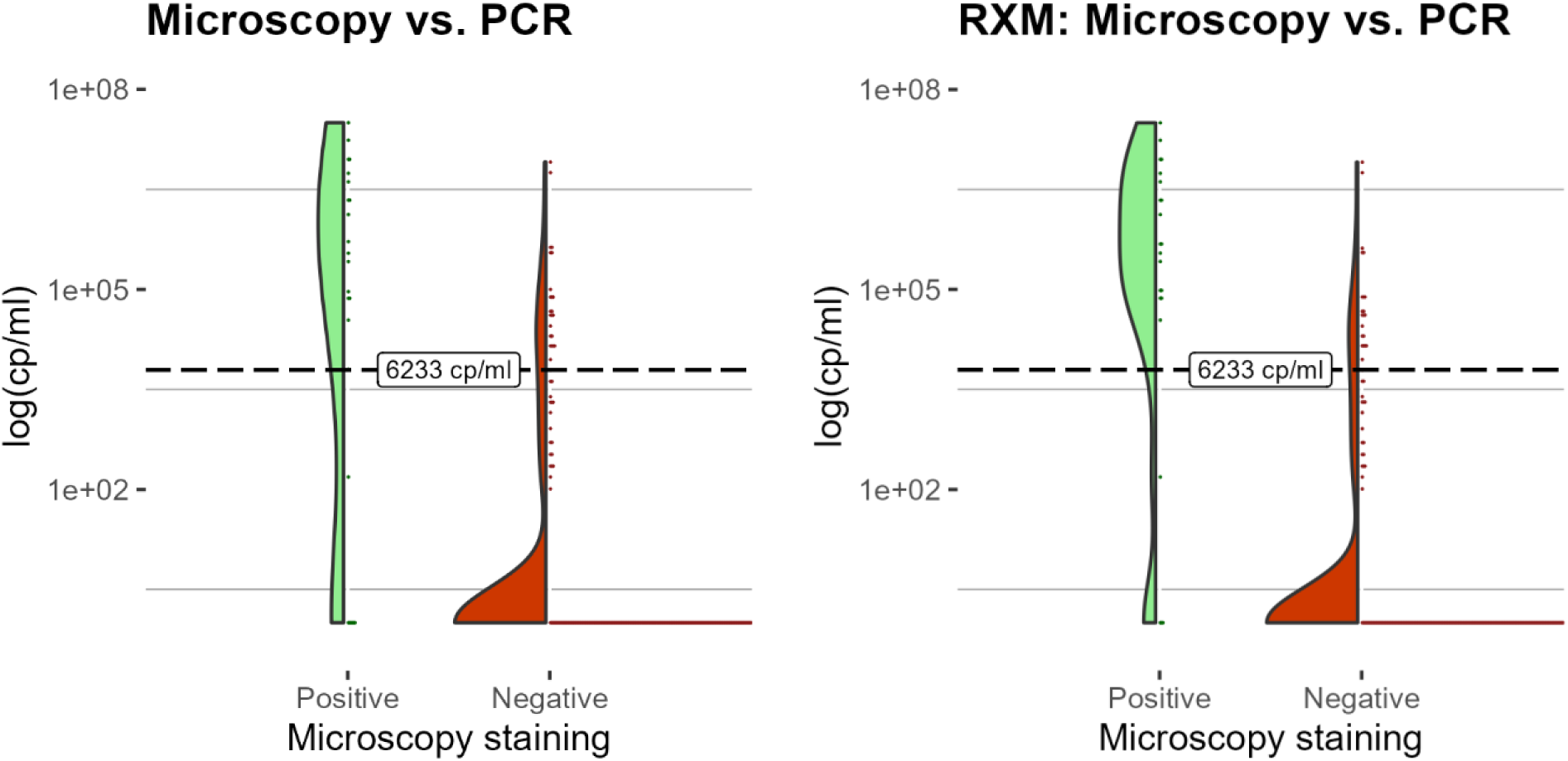
Distribution of PCR copy numbers (log10 scale) stratified by microscopy result with a dashed line indicating the optimal cut-off at 6233 cp/ml based on the Youden index. **a)** Primary data; **b)** Reexamination (RXM)

Sensitivity and specificity vary depending on the selected cp/ml cut-off, as does the Youden index incorporating both parameters (Sensitivity + Specificity −1). As shown in Figure 4, specificity increases steadily with higher thresholds, while sensitivity declines more rapidly beyond a certain point. The Youden index peaks at a cut-off of 6233 cp/ml, representing optimal balance between sensitivity and specificity in both the “Primary data” and “Reexamination” datasets.

**Figure 4:**
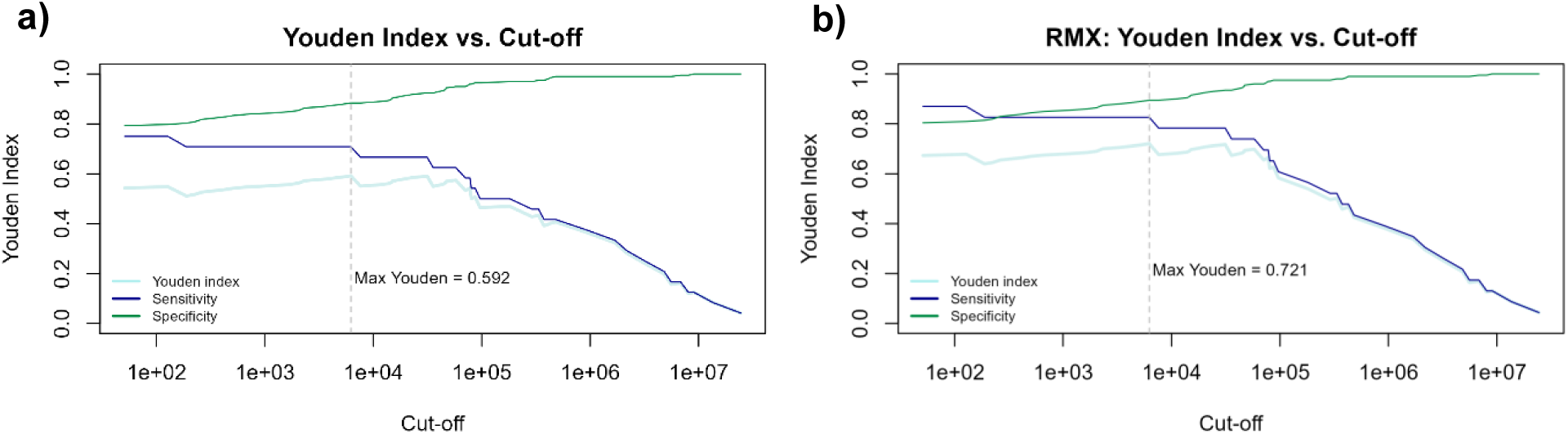
Youden index, sensitivity, and specificity plotted against PCR thresholds with the optimal cut-off (6233 cp/ml) indicated as dashed line by peak Youden index (Max Youden). **a)** Primary data; **b)** Reexamination (RXM)

### Subgroup analysis of IF staining method and sample type

The calculated performance of the PCR assay compared to the reference standard varied depending on the IF staining method and sample type. IF assays from two different suppliers were used: Bio-Rad (*n*=148) and Merifluor (*n*=74). Samples stained by Bio-Rad demonstrated a better performance, with AUC values of 0.92 (Primary Data) and 0.98 (Reexamination), compared to Merifluor-stained samples (supplementary Figure S1 and Figure 2). This was also reflected in the Bio-Rad group’s sensitivity and specificity at the optimal threshold of 6233 cp/ml in both datasets (Table 2). Sensitivity and specificity reached 88.9% and 92.8%, respectively, and improved to 100% and 92.9% after reexamination. It is worth mentioning that at its optimal threshold only the Bio-Rad subgroup achieved zero false negatives and 100% sensitivity after the reexamination.

**Table 2:** Diagnostic performance of PCR across datasets and subgroups. Optimal cut-offs were calculated using Youden’s index (Sensitivity + Specificity - 1).

|  | AUC | Cut-off (cp/ml) | Sensitivity | Specificity |
| --- | --- | --- | --- | --- |
| **Primary Data** |  |  |  |  |
| All samples | **0.82** | **6233** | **0.71** | **0.88** |
| BR | 0.92 | 6233 | 0.89 | 0.93 |
| Mf | 0.73 | 181178 | 0.53 | 0.92 |
| BAL | 0.87 | 5298.5 | 0.78 | 0.90 |
| SP | 0.68 | 4198338 | 0.50 | 0.98 |
| **Reexamination** |  |  |  |  |
| All samples | **0.90** | **6233** | **0.83** | **0.90** |
| BR | 0.98 | 6233 | 1 | 0.93 |
| Mf | 0.82 | 57749.5 | 0.73 | 0.87 |
| BAL | 0.94 | 5298.5 | 0.89 | 0.92 |
| SP | 0.75 | 4198338 | 0.60 | 0.98 |
| **Clinical reevaluation\*** |  |  |  |  |
| All samples | **0.92** | **31377** | **0.81** | **0.95** |
| **No false negatives\*** |  |  |  |  |
| All samples | **0.98** | **6233** | **0.96** | **0.91** |

\*Two additional datasets were used for sensitivity analysis: "Clinical reevaluation"
reclassified patients with confirmed PJP as true positives, while "No false negatives" further excluded all potential false-negative cases (see section "Sensitivity analysis").
Abbreviations: AUC, area under the curve; BR, Bio-Rad; Mf, Merifluor; SP, sputum

In contrast, samples stained by Merifluor required higher thresholds to optimize diagnostic performance (181’178 before and 57’749.5 after reexamination). Even in the “Reexamination” dataset, sensitivity and specificity reached only 73.3% and 88.1%, respectively, compared to 53.3% and 91.5% in the “Primary Data”.

Sample type also influenced assay performance. Among the 222 samples, 152 were BAL and 70 were sputa. BAL samples showed higher AUC values (0.87 and 0.94 after the reexamination) than sputum samples (0.68 and 0.75 after the reexamination) (Figure 2, Table 2).

### Evaluation of patient characteristics from discrepant results

We reviewed patient histories for all six IF-positive/PCR-negative cases and 10 of the 13 IF-negative/PCR-positive samples with high PCR positivity (Table 3 and detailed clinical descriptions in supplementary Table S1).

**Table 3:**
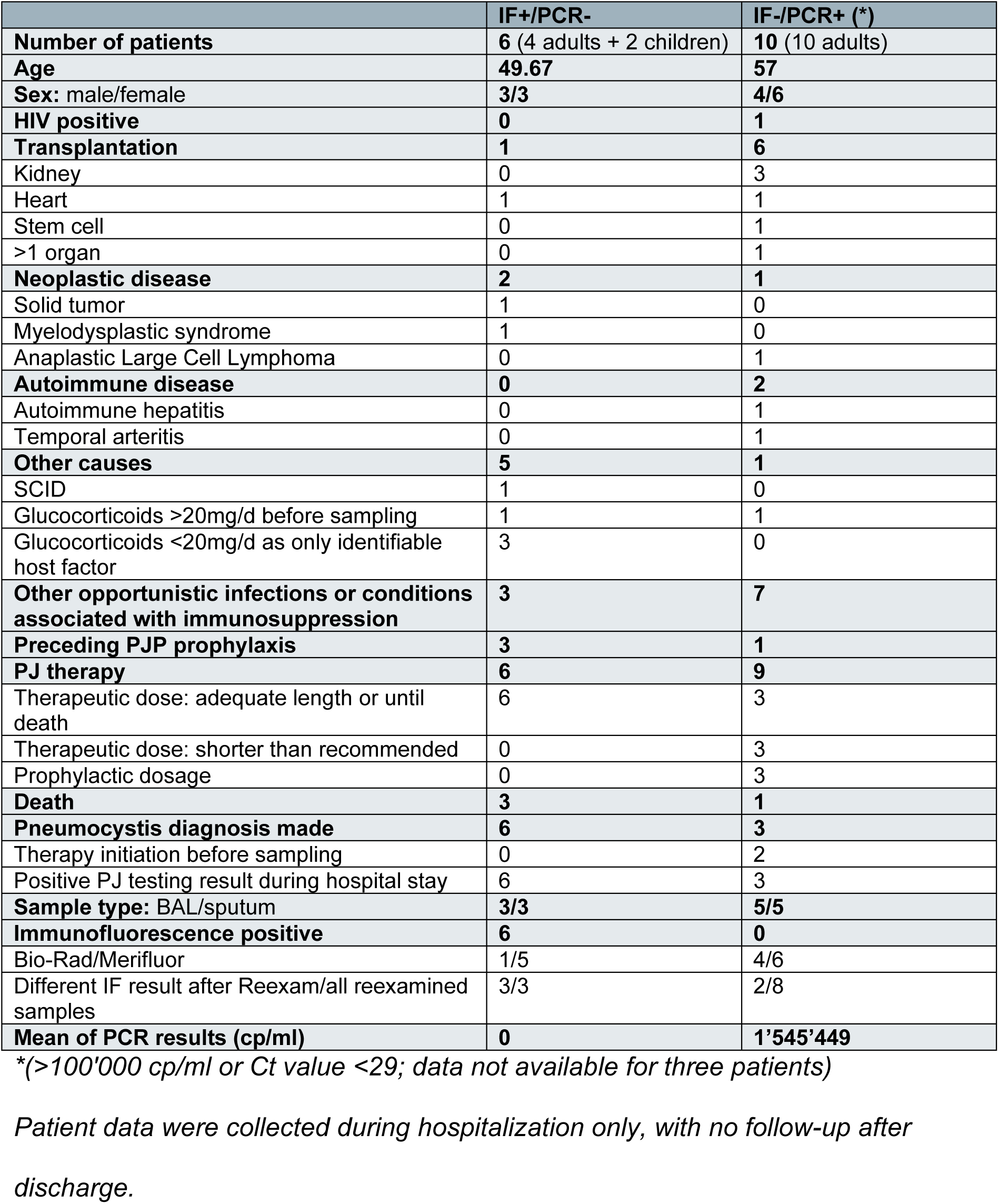
Clinical characteristics of patients with discrepant IF/PCR results. Left: IF+/PCR− cases; Right: IF−/PCR+ cases with high PCR positivity.

Both subgroups included a similar mix of BAL and sputum samples. However, in the IF-positive/PCR-negative group, five of six samples were stained with Merifluor rather than Bio-Rad, including all that could not be reexamined by IF (*n*=3/3). In three of the six cases, low-dose glucocorticoid use (≤15 mg/d) was the only identifiable host factor, which is generally not considered clinically relevant^34^. All six IF-positive/PCR-negative patients received therapeutic doses of trimethoprim-sulfamethoxazole (TMP-SMX), with three already on prophylaxis. The impact of TMP-SMX remains unclear in 3/6 cases due to their death within the first 10 days following PJP diagnosis.

In the IF-negative/PCR-positive subgroup (*n*=13), despite negative IF results, three patients received prophylactic TMP-SMX and six received full treatment. In three of these treated patients, therapy was stopped within six of the recommended 21 days^35^. The other three completed ≥21 days of therapy and were formally diagnosed with PJP based on a later IF-positive BAL after an initial negative sputum in one patient; a highly positive external PCR result in another patient; and a prior IF-positive result (13 days earlier) with treatment initiation in the third patient. Two other IF-negative/PCR-positive patients had recent or ongoing PJ prophylaxis.

Based on these findings, additional data processing for a subsequent sensitivity analysis was conducted. First, a dataset named “Clinical reevaluation” was created, in which the three IF-negative/PCR-positive patients with confirmed PJP diagnosis during hospitalization were reclassified as true-positive, due to the strong clinical and diagnostic evidence (supplementary Figure S2). Using the assay’s limit of detection (LoD; 97 cp/ml) as the cut-off, sensitivity increased to 88.5%, specificity to 74.4% and the AUC reached 0.92 in this dataset. The optimal threshold was identified at 31’377 cp/ml with a sensitivity of 80.8% and specificity of 94.9% (Table 2).

As the three IF-positive/PCR-negative samples that we could reexamine proved to be true-negatives, we reclassified the three remaining IF-positive/PCR-negative samples true-negative in a final sensitivity analysis (supplementary Figure S3). This dataset referred to as “No false negatives”, reached a sensitivity of 100% and a specificity 75% (supplementary Figure S2). The optimal threshold remained at 6233 cp/ml consistent with the “Primary data” and “Reexamination” datasets (Table 2) achieving a sensitivity of 95.7% and specificity of 91%. To reach a sensitivity of 100%, the threshold would have to be lowered to 128 cp/ml with a corresponding drop in specificity to 82.4%.

## Discussion

In our study, IF detected PJ in 24 samples whereas PCR identified 74 positive samples. Based on these findings, we performed ROC analyses and determined diagnostic thresholds using Youden’s index. Although IF was used as the reference standard, it is important to note that no universally accepted gold standard exists for identifying an active PJ infection^36–38^. Thus, we conducted reexaminations of discrepant samples using IF, reviewed patient histories of discrepant samples, and performed a sensitivity analysis excluding false-negative results to contextualize our results. Our findings are in line with previous reports showing that quantitative PCR is more sensitive than IF for detecting PJ, supporting its implementation in the diagnostic process of PJP^20, 36–51^.

Direct pathogen detection is a crucial part in the diagnosis of PJP and should be interpreted alongside the pretest probability from clinical and radiological findings^2, 4^. In recent years, several PCR assays have been developed to replace microscopic detection in routine diagnostics^19^. Our findings reflect the reported advantages of PCR, including higher sensitivity, reduced observer-dependency, and improved time-efficiency^9, 19, 23^.

Among the 62 discrepant cases, 56 were IF-negative but PCR-positive. 13 showed a highly positive PCR result (>100’000 cp/ml or Ct value <29) and eight of them had enough material for IF reexamination with PJ detected in two. Clinical reevaluation showed that three additional cases were diagnosed with PJP during hospitalization, including two with negative reexaminations and one lacking material, further supporting the higher sensitivity of PCR.

In the first case, the original IF report noted “questionable trophozoites found; re-sampling suggested” and a follow-up BAL two days later tested IF-positive. The second patient, previously diagnosed and treated for PJP, had a negative IF 13 days later but remained PCR-positive (318,229 cp/ml), likely reflecting residual DNA fragments indicating a limitation of PCR in monitoring short-time treatment response. A third HIV-positive patient showed IF-negative results one day after treatment initiation but tested strongly positive by both external PCR (2,000,000 cp/ml) and our assay (5,644,021 cp/ml).

Although the remaining IF-negative/PCR-positive cases were not formally diagnosed with PJP, many received TMP-SMX either therapeutically or prophylactically. Active PJ infection or colonization could not be definitively excluded.

The IF-positive/PCR-negative group included six non-HIV patients. Three samples were reexamined, and none confirmed the initial IF result, suggesting true negatives. One had a prior note indicating the observed cyst could not be verified by a second examiner. The remaining three samples, all stained with Merifluor, were associated with glucocorticoid use, though two of them received only a low dosage (<20mg/d) as the only identifiable host factor.

Clinical context supports the possibility of false-positive initial IF results. One patient on 15 mg/day prednisolone for lymphocytic pneumonia and ongoing prophylaxis, died shortly after IF positivity and TMP-SMX initiation, preventing assessment of treatment response. Persistent trophozoites from prior infection even after successful treatment may explain the positive IF^37, 52^. Another patient with COPD GOLD3, using intermittent inhaled steroids, improved on TMP-SMX, though alternative pathogens could not be excluded. The third patient started glucocorticoid treatment (60mg/d) three days before sample collection, while suffering from multi-organ failure and E.coli sepsis. This patient died 10 days later and the impact of TMP-SMX remains unclear.

In the literature, 13 of 17 studies comparing IF and PCR reported no IF-positive/PCR-negative cases^20, 36, 39–43, 45, 47–51^. Of the remaining four studies, one did not perform IF on PCR-negative samples, and three reported IF-positive/PCR-negative results^37, 38, 44, 46^. In one study, four such cases were observed among 156 IF-positive samples (305 total samples total, 204 PCR-positive) attributed to two PCR inhibitors, one false-positive IF result and one unidentified technical error^38^. Another study reported five IF-positive/PCR-negative results out of seven IF-positives (222 total samples, 93 PCR-positive) with one linked to radiation pneumonitis^37^. The third study documented one IF-positive/PCR-negative case among 33 IF-positive samples (411 total samples, 61 PCR-positive), without specifying the cause^44^. Definitions of IF positivity also vary across studies and while our study considered any visible PJ form (trophozoites, cysts) as positive, others required ≥2 or ≥5 cysts for a positive result^37, 38, 45, 46^. Given these factors, the cause of IF-positive/PCR-negative discrepancies remains uncertain. Potential explanations include Merifluor-related false positive results, residual organisms post-treatment, observer variability or sample degradation. PCR inhibition can be ruled out due to the assay including an internal control, making this explanation unlikely.

The PCR assay’s limit of detection (97copies/ml) is not appropriate as a diagnostic threshold in clinical practice. Using it as a cut-off may lead to unnecessary treatment, particularly in patient populations with a potentially higher susceptibility to colonization, such as children, immunosuppressed adults, individuals with chronic lung diseases, and healthcare workers^53^. While specificity is important, ensuring high sensitivity is crucial given the high mortality associated with PJP. Therefore, identifying a clinically meaningful cut-off that balances both sensitivity and specificity is essential for accurate diagnosis and appropriate patient management. In our study, the optimal threshold, based on the highest Youden index, was consistently identified as 6233 cp/ml across the “Primary data”, “Reexamination” and “No false negatives” datasets. An optimal cut-off value ideally discriminates between an acute infection and colonization.

One study aiming to make this distinction and targeting the mtLSU rRNA gene proposed similar thresholds, suggesting <10^3^ cp/ml may indicate colonization and >5×10^3^ cp/ml an acute infection^54^. Another study using the same PCR target reported 100% sensitivity <1.6×10³ cp/ml and 100% specificity >2×10⁴ cp/ml for active PJ infection^55^. Other authors with PCR assays targeting other PJ-specific genes proposed cut-offs of >1.3×10³ cp/ml and >1.45×10³ cp/ml for a high probability of PJP^36, 56^. Although cut-off values are assay-specific, our findings align with other previously published thresholds and thus, our analysis suggests the following diagnostic algorithm for PJP identification using PCR results (Figure 5). The cut-off values are derived from the validation of our specific PCR assay while integrating previous literature and its validity relies on accurate pre-analytical processing.

**Figure 5:**
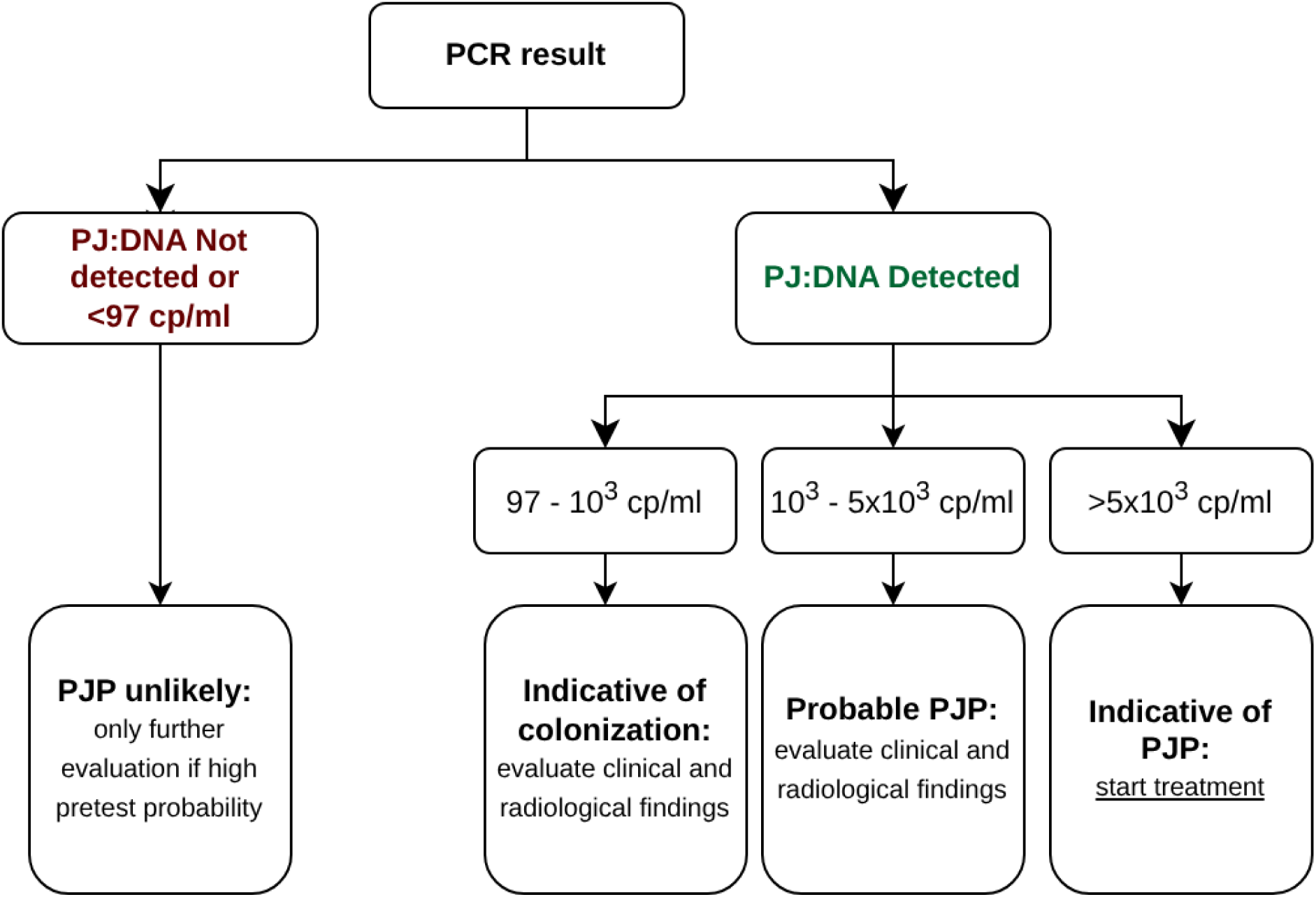
Flowchart illustrating the PJP diagnostic algorithm based on PCR results, highlighting the clinical consequence of different cp/ml cut-off values.

A sensitivity of 100% was achieved only after reexamination in the Bio-Rad group at the 6233 cp/mL threshold, and in the “No false negatives” dataset at a lower threshold of 128 cp/ml. Importantly, the used IF assay affects only the reference standard, not the PCR itself. The potentially lower specificity of Merifluor likely influenced the apparent performance of PCR when all samples are considered. 83.3% of IF-positive/PCR-negative results were analyzed using Merifluor, including all three that could not be reexamined. The reexamination was conducted using Bio-Rad. Literature suggests Merifluor may have higher sensitivity than conventional microscopic stains but also lower specificity with a significantly higher false-positive rate^31–33^. One author describes a positive predictive value (PPV) of 81.9% for Merifluor, compared to >96% for the other conventional microscopic stains (Diff-Quik, Grocott-Gomori methenamine silver (GMS), and Calcofluor white stains)^31^. Another study reports one IF-positive/PCR-negative result from a total of 83 tested samples^33^.

The broad gap between thresholds achieving 100% sensitivity and those yielding 100% specificity illustrates the sensitivity-specificity trade-off, introducing diagnostic uncertainty in interpreting intermediate PCR results. This reinforces the importance of integrating pretest probability from clinical and radiological findings into diagnostic decision-making. In some cases, treatment may be warranted even without any direct pathogen detection. While PCR’s high sensitivity may lead to false positives due to its inability to differentiate colonization from active infection, the risk of missing a true PJP diagnosis cannot be ignored^23, 24^.

A major issue in evaluating PJ PCR assays is the absence of a universally accepted gold standard for PJP diagnosis even though IF remains widely regarded as the current standard method^36–38^. Both PCR and microscopy are influenced by sample quality and type, but IF is further affected by the utilized assay. Our findings suggest that Merifluor may reduce IF specificity and representing a potential bias in PCR performance analysis. Additionally, prior treatment may degrade fungal structures and disproportionately affecting IF sensitivity. The predominant presence of trophozoites during active infection may also negatively impact microscopic detection^23^. These factors highlight the limitations of using IF as a reference standard and justify our sensitivity analyses, which demonstrated excellent performance of the ELITe InGenius PJ PCR assay. Additionally, all specimens except non-mucoid BAL were diluted during IF preparation and stored frozen for at least eight months prior to analysis. Of the 222 samples examined, only 24 tested positive by IF, meaning that a small number of cases disproportionately influenced sensitivity and specificity estimates. Finally, the absence of residual material in certain cases limited the ability to perform IF reexaminations and the reliance on pre-existing data inherent to the retrospective design represented additional limitations.

In conclusion, our data suggests a higher sensitivity of the ELITe InGenius PJ PCR assay compared to IF for direct PJ detection. Confirmed PJP diagnoses in IF-negative/PCR-positive cases support this finding. Although the origin of discrepant results cannot always be definitively determined, the 100% negative reexamination among IF-positive/PCR-negative samples and the known limitations of IF support the reliable performance of PCR. Overall, given its higher sensitivity and robust performance, the ELITe InGenius PJ PCR assay represents a valuable addition to routine diagnostics for PJP with the potential to improve early detection and guide timely treatment decisions.

## Data Availability

All data produced in the present work are contained in the manuscript

## Acknowledgments

We thank Noelia Berther for excellent technical assistance. This work was supported by the Swiss National Science Foundation (grant no. 211422) to SDB.

## Supplemental material

**Figure S1:**
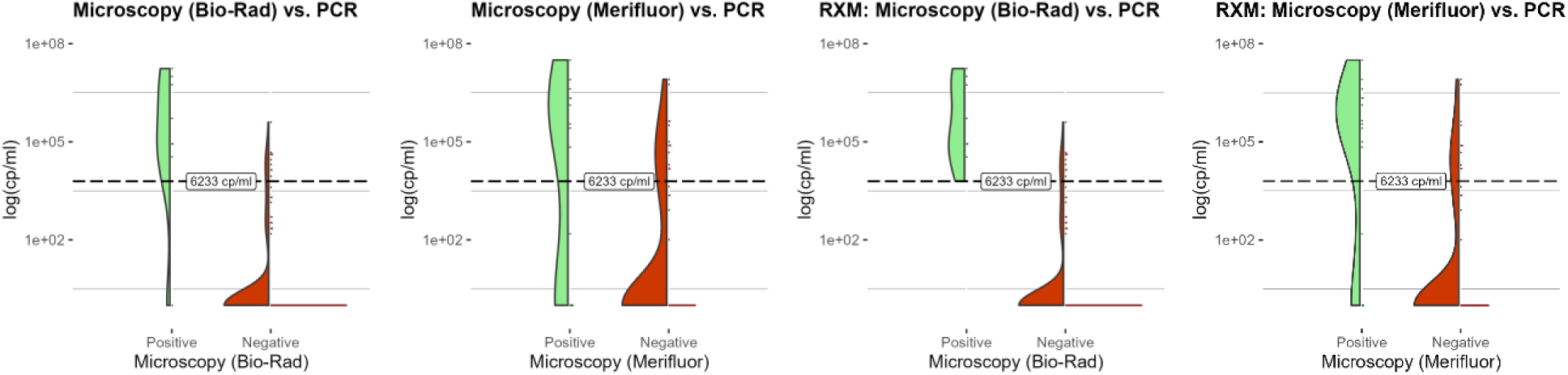
PCR copy number distribution (log10 scale) stratified by IF assay supplier. **a)** Primary data **b)** Reexamination (RXM)

**Figure S2:**
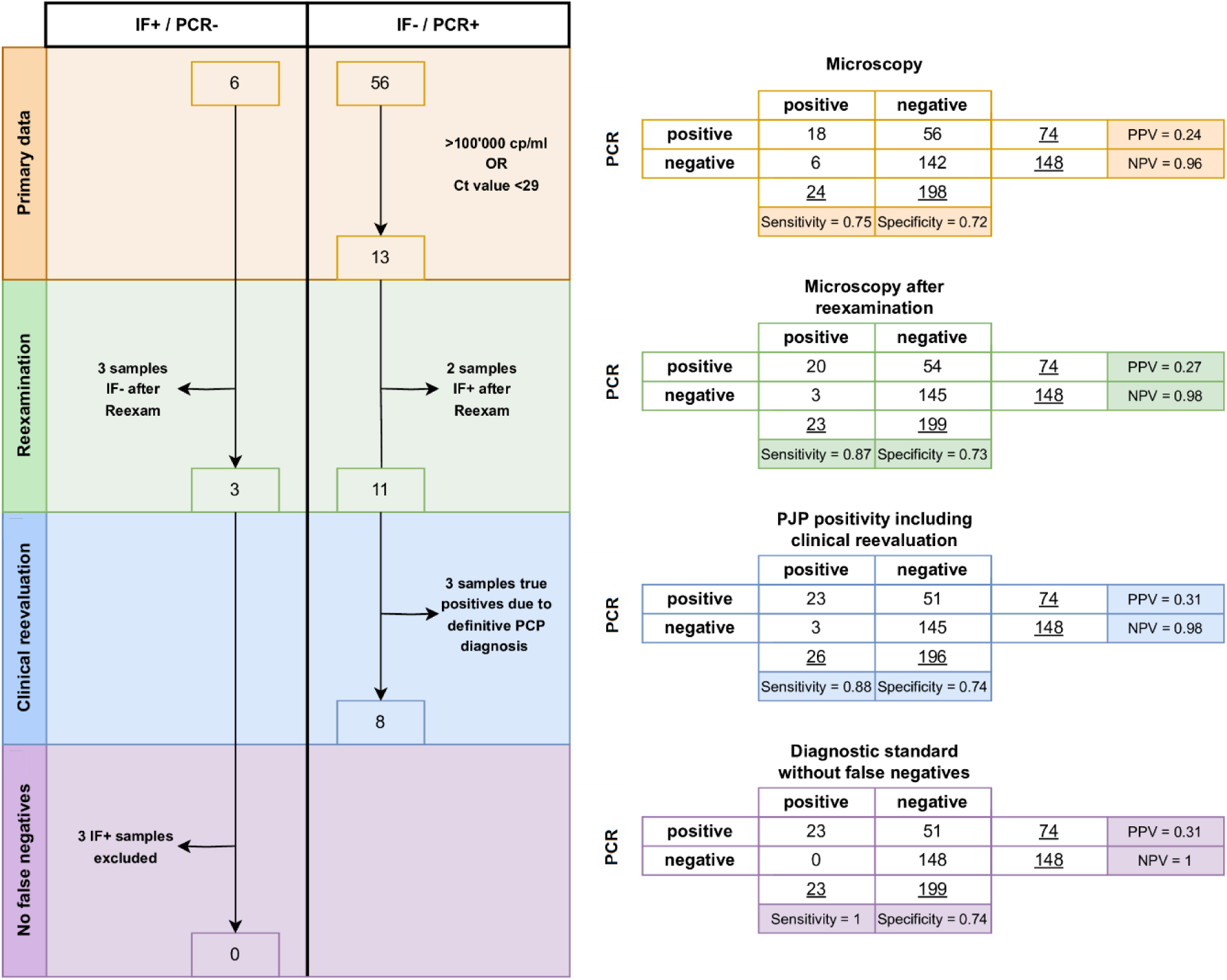
Flowchart showing data processing of discrepant results in microscopy and PCR analysis. The unadjusted primary data (orange) includes six IF+/PCR- and 56 IF-/PCR+ samples, 13 of which had high PCR positivity. IF reexamination (green) was performed in three IF+/PCR- and eight IF-/PCR+ cases and revealed three true-negative and two true-positive samples. The ‘Clinical reevaluation’ (blue) revealed further three confirmed PJP cases with negative IF. The final dataset, ‘No false negatives’ (violet), further removes the remaining three false-negative PCR cases.

**Figure S3:**
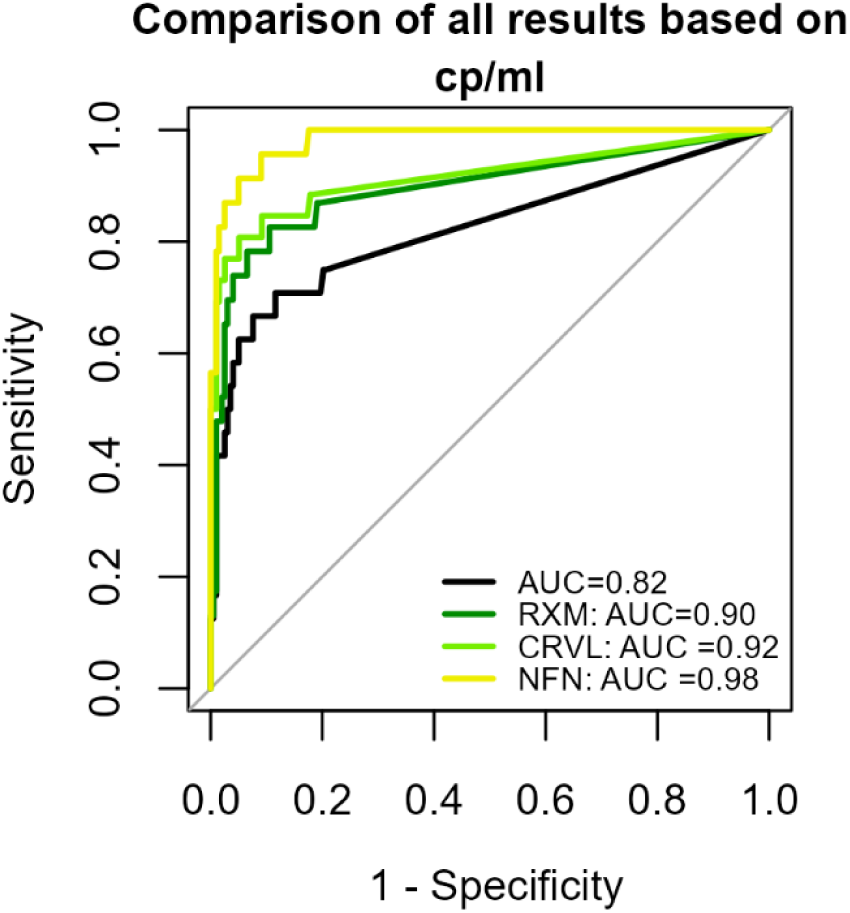
ROC curves comparing PCR performance across four datasets: “Primary data”, “Reexamination” (RXM), Clinical reevaluation (CRVL), and “No false negatives” (NFN).

**Table S1:** Detailed clinical information of patients with IF+/PCR− and IF−/PCR+ results.

**a) IF+/PCR–**
| Re | ST | IFA | Host factor | Prophx | PJP therapy | Clinical course;<br>treatment response | Additional Info |
| --- | --- | --- | --- | --- | --- | --- | --- |
| **Possible or probable PJP** |  |  |  |  |  |  |  |
| NA | SP | Mf | <u>GC inh. if required due to COPD GOLD 3; MDS (unlikely host factor per oncologists)</u> | - | **th:TMP-SMX** | Suitable clinical presentation and radiology; adequate | Oral candidiasis (Fluconazole treatment), HSV1-reactivation |
| **Unclear** |  |  |  |  |  |  |  |
| neg | BAL | BR | <u>SCID (Severe Combined Immunodeficiency)</u> | Bactrim | **th:TMP-SMX** | Suitable clinical presentation; unclear | 2d post-SC after revisiting the same sample: only cyst not found anymore |
| neg | BAL | Mf | <u>s/p heart-TPL: MMF, Tac, Ever</u> | Nopil | **th:TMP-SMX** | Suitable clinical presentation; adequate | Poor treatment response to Co-Amoxicillin |
| NA | SP | Mf | <u>Pred p.o. 15mg/d due to lymphocytic pneumonia</u> | Bactrim | **th:TMP-SMX**<br>1d until death | Suitable clinical presentation and radiology; unclear (death: 2d post-SC) | St.n. *Pneumocystis jirovecii* pneumonia, ca. 90d pre-SC (PCR +) |
| NA | BAL | Mf | <u>GC i.v. 60mg/d from 3d-preSC – death; severe underlying multisystem disease</u> | - | **th:TMP-SMX**<br>10d until death | Decompensated cardiac insufficiency + congestive and chronic hepatopathy; unclear (death: 10d post-SC, septic and hemorrhagic shock ) | *E.coli* (4/4): BC (16d pre-SC), *C. dubliniensis*: UC (11d pre-SC) + BAL (present sample), Yeast fungus: UC (5d post-SC) |
| **Alternative diagnosis** |  |  |  |  |  |  |  |
| neg | SP | Mf | <u>Pred p.o. (≤10mg/d) if required due to COPD; SCLC (limited disease, no treatment)</u> | - | **th:TMP-SMX**<br>7d until death | Suitable clinical presentation, radiologically unclear; poor (death 7d post-SC) | *S. aureus* found in the sputum sample, additional to PJ |

**b) IF-/PCR+ (>100'000 cp/ml or Ct value <29; data not available for three patients)**
| Re | ST | IFA | Host factor | Prophx | PJP therapy | Clinical + Outcome | Additional Info |
| --- | --- | --- | --- | --- | --- | --- | --- |
| **Definite PJ diagnosis** |  |  |  |  |  |  |  |
| neg | BAL | Mf | <u>s/p kidney-TPL</u>: Tac, MMF, Pred, induction anti-T-Lyc-globulin-Ig | - | **th:TMP-SMX**<br>6d: from 13d–7d pre-SC;<br>**Clinda + PQ**<br>7d pre-SC–10d post-SC; afterwards Atovaq (pr) | Definite PJP diagnosis (13d pre-SC), 16d pre-SC suitable radiology and improvement seen on day of sample collection | Positive with IFA + TOL + PCR;<br>*Aspergillus*: BAL (13d pre-SC),<br>*P. aeruginosa*: TS (16d pre-SC) |
| NA | SP | Mf | <u>s/p combined kidney- and liver-TPL</u>: Tac, MMF, Pred 5mg, induction BXM | - | **th:TMP-SMX**<br>21d: SC–21d post-SC | Definite PJP diagnosis (2d post-SC), suitable radiology; 10d post-SC clinical improvement | Commentary: “questionable trophozoites found; re-sampling suggested”; 2d post-SC: Follow-up BAL sample IFA positive; Pred 40mg |
| neg | BAL | Mf | <u>HIV1 C3</u> with opportunistic infections due to malcompliance; (HIV PCR = 405'000 cp/ml, CD4 = 183 cells/μl) | - | **th:TMP-SMX**<br>21d: 1d pre-SC–20d post-SC | Definite PJP diagnosis (1d pre-SC), suitable radiology; later clinical improvement | Fluconazole (6d–9d post-SC for oral candidiasis), Pred (1d pre-SC–20d post-SC); External PCR (2*10<sup>6</sup> cp/ml) 3d post-SC, follow-up IF with new staining still negative |
| **Possible or probable PJP** |  |  |  |  |  |  |  |
| neg | SP | BR | <u>Large T cell lymphoma</u>: MPred 80mg (1d pre-SC) | - | **th:TMP-SMX**<br>5d: SC–5d post-SC; afterwards prophylactic | Lower abdominal pain at admission; development of type 1 respiratory failure, 5d post-SC clinical improvement, 7d post-SC develops fever again; 19d post-SC discharge | 1d pre-SC: MPred 80mg d1 + 160mg d2<br>From SC: MPred 100mg, ETO, CDV, G-CSF; HSV1-Reaktivierung (7d post-SC),<br>E. faecium: UC (7d post-SC) |
| pos | BAL | Mf | <u>s/p stem cell-TPL</u>: acute fibrinous and organizing pneumonia most likely due to GvHD II | Bactrim, until 90d pre-SC | **pr:TMP-SMX**<br>7d post-SC–while high-dose GC therapy | Suitable clinical presentation and radiology, LDH↑; 8d post-SC clinical improvement | MPred i.v. 125mg (4d–7d post-SC),<br>Pred 120mg (7d post-SC–XX);<br>Adenovirus: PCR + |
| neg | BAL | Mf | <u>s/p heart-TPL</u>: MMF, Tac, Ever, Pred 2.5mg, induction BXM; dialysis-dependent CKD | - | **th:TMP-SMX**<br>6d: 1d pre-SC–5d post-SC | Suitable clinical presentation and radiology; 11d post-SC clinical improvement | Ceftriaxon 6d–1d pre-SC |
| **Alternative diagnosis** |  |  |  |  |  |  |  |
| NA | SP | BR | <u>GCA</u>: Pred p.o. 60mg/d (46d pre-SC) - 20mg (ca. 30d post-SC); Gout: Pred p.o. 50mg if required; dialysis-dependent CKD | Bactrim | **pr:TMP-SMX**<br>1d pre-SC–21d post-SC | Suspected COVID-19 pneumonia with possible bacterial superinfection; 10d post-SC clinical improvement | Cefepim 1g (1d pre-SC), Nirmatrelvir/Ritonavir (1d pre-SC–3d post-SC), Metronidazol (SC–3d post-SC) |
| neg | BAL | Mf | <u>s/p kidney-TPL</u>: CyA, MMF, dialysis-dependent CKD | - | **pr:TMP-SMX**<br>9d post-SC–while high-dose GC therapy | Cryptogenic organizing pneumonia (ED 7d post-SC); 9d post-SC clinical improvement | Co-Amoxicillin (1d pre-SC – 13d post-SC),<br>Pred 30mg + Amphotericin B (11d post-SC–XX) |
| **Unclear** |  |  |  |  |  |  |  |
| NA | SP | BR | <u>Autoimmune hepatitis</u>: MPred i.v. 100mg (13d pre-SC–7d pre-SC), Pred p.o. 75mg (6d pre-SC–death), Aza (6d pre-SC–death) | - | - | Unclear (acute Hepatitis, respiratory insufficiency); unclear (death: 4d post-SC, septic shock) | *S. aureus*: BC (4/4) (2d post-SC), UC+BC (1/4) (SC);<br>*A. fumigatus*: SP (1d post-SC);<br>Vancomycin i.v. (3d pre-SC), Piperacillin/Tazobactam (3d pre-SC – death) |
| NA | SP | BR | s/p heart-TPL: Tac,<br>MMF, Pred 5mg | - | th:TMP-SMX<br>2d: 5d–7d<br>post-SC | Decreasing kidney<br>function, Suitable<br>clinical presentation<br>and radiology, LDH↑;<br>5d post-SC pulmonary<br>deterioration; 11d<br>post-SC clinical<br>improvement | 3 additional samples (IFA-<br>/PCR+): 2x SP (ca. 1d post-<br>SC), BAL (7d post-SC);<br>Meropenem (6d–10d post-<br>SC),<br>Levofloxacin (1d–3d post-<br>SC + 6d–14d post-SC),<br>Co-Amoxicillin (9d–14d post-<br>SC) |

*All sample collections took place in 2023 besides one marked with (\*). All dates mentioned in one bracket refer to the same year as the sample collection.*
*Abbreviations: Re, result after Reexam; ST, sample type; IFA, IF assay; Prophx, PJP prophylaxis; d, day(s); SC, day of sample collection; pre-SC, before sample collection; post-SC, after sample collection; s/p, status post; inh., inhalative; p.o., peroral; i.v., intravenous; th, therapeutic; pr, prophylactic; PQ, Primaquine; Clinda, Clindamycin; GC, Glucocorticoids; Pred, Prednisone; MPred, Methylprednisone; Aza, Azathioprine; CyA, Cyclosporin A; Tac, Tacrolimus; MMF, Mycophenolat mofetil; BXM, Basiliximab; ETO, Etoposid; CDV, Cyclophosphamid + Doxorubicin + Vincristin; Ever, Everolimus; d., death; BC, blood culture; UC, urine culture; TS, tracheal secretion; CDK, chronic kidney disease, GCA, giant cell arteritis; XX, unknown date*

## Notes

### Competing Interest Statement

The authors have declared no competing interest.

### Author Declarations

The ethics committee of the Canton Zurich reviewed the study protocol (Kantonale Ethikkommission Zurich BASEC ID Req-2025-01064) and concluded that the work did not meet the criteria for requiring ethical approval within its remit.

